# Association Between Motor Function and Higher-Order Repetitive Behaviors in Monogenic Autism Spectrum Disorder

**DOI:** 10.64898/2026.01.21.26344541

**Authors:** Shelby Smout, Seulgi Jung, Aishwaryaa Udeshi, Madison Caballero, Amy Rapp, Alexander Kolevzon, Behrang Mahjani

## Abstract

**Importance:** Motor skill impairments affect up to 87% of children with autism spectrum disorder (ASD) and are associated with greater severity of repetitive behaviors. Yet, most research examining this relationship has treated ASD as a unitary condition. Understanding whether motor-behavior relationships differ by genetic etiology could inform stratified approaches to ASD research and clinical care.

**Objective:** To determine whether the relationship between motor function and restricted and repetitive behaviors (RRBs) differs between children with monogenic forms of ASD (*SHANK3, DYRK1A*, or *SCN2A* variants) and children with idiopathic ASD.

**Design, Setting, and Participants:** Matched cohort cross-sectional study using data from the Simons Foundation Powering Autism Research for Knowledge (SPARK) database. Children with loss-of-function variants in *SHANK3, DYRK1A*, or *SCN2A* were matched to children with idiopathic autism and intellectual disability.

**Main Outcomes and Measures:** Motor function was assessed using the Developmental Coordination Disorder Questionnaire (DCDQ). Repetitive behaviors were assessed using the Repetitive Behavior Scale-Revised (RBS-R), with subscales categorized as lower-order (stereotyped, self-injurious) or higher-order (compulsive, ritualistic, sameness, restricted interests). The primary analysis compared motor-RRB correlations between groups.

**Results:** The sample included 93 children with monogenic autism (*SHANK3*, n=34; *DYRK1A*, n=46; *SCN2A*, n=13) and 787 matched children with idiopathic ASD. In idiopathic ASD, motor function was negatively correlated with RRBs (r = -0.156); in monogenic ASD, this reversed to a positive correlation (r = +0.185; Δr = 0.341, P = 0.002). This reversal was specific to higher-order RRBs (idiopathic r=-0.106; monogenic r=+0.234; Δr=0.339, 95% CI 0.124-0.535, P=0.002) and was not observed for lower-order RRBs (Δr=0.212, P=0.05). All three genes showed positive correlations (*SHANK3* r=+0.033; *DYRK1A* r=+0.262; *SCN2A* r=+0.623) with no significant heterogeneity (P=0.153).

**Conclusions and Relevance:** The relationship between motor function and repetitive behaviors differs by genetic etiology, with children with monogenic ASD showing a positive motor-RRB correlation specific to higher-order behaviors, opposite to the negative correlation observed in idiopathic ASD. This reversal was consistent across three molecularly distinct genes. These findings support stratifying autism research and clinical care by genetic etiology.

**KEY POINTS:** *Question:* Does the relationship between motor function and restricted and repetitive behaviors (RRBs) differ between children with autism spectrum disorder (ASD) attributable to *SHANK3, DYRK1A*, or *SCN2A* variants and children with idiopathic ASD?

*Findings:* We conducted a matched cohort cross-sectional study comparing correlations between motor function and RRBs in children with monogenic ASD versus children with idiopathic ASD and intellectual disability. Motor function was negatively correlated with RRBs in children with idiopathic ASD but positively correlated in children with monogenic ASD.

*Meaning:* Genetic variants may alter behavioral organization, supporting the value of stratifying populations of individuals with ASD by genetic etiology in both research and clinical care.

## INTRODUCTION

Autism spectrum disorder (ASD) affects approximately 1 in 31 children in the United States and is defined by two core symptom domains: deficits in social communication and restricted, repetitive behaviors (RRBs).^1,2^ However, ASD encompasses substantial variability in clinical presentation, developmental trajectory, and underlying biology.^3,4^ This heterogeneity extends across cognitive profiles, adaptive behavior, neuroanatomical features, and the relationships between behavioral domains, raising the question of whether different etiological forms of ASD share the same behavioral architecture or differ in fundamental ways.^5^

Motor skill impairments, while not a diagnostic criterion, are among the most common co-occurring features of ASD, affecting up to 87% of individuals with ASD.^6-8^ Large-scale analyses of the Simons Foundation Powering Autism Research for Knowledge (SPARK) cohort have demonstrated that risk for motor impairments is over 22 times greater in children with ASD compared to the general population, with motor difficulties persisting throughout childhood and predicting both social communication deficits and repetitive behavior severity even after accounting for intellectual disability.^8,9^ A growing body of research has documented associations between motor function and RRBs in ASD, with better motor coordination generally associated with fewer RRBs.^10-12^ This relationship has been attributed to shared neural substrates, particularly frontal-striatal-cerebellar circuits that support both motor control and behavioral regulation.^13-15^ Structural and functional neuroimaging studies have demonstrated that sensorimotor regions of the cerebellum and their cortical connections show abnormalities that correlate with both motor deficits and increased repetitive and stereotyped behaviors.^14,16^ Importantly, these circuits support not only motor coordination but also cognitive flexibility and error-based learning, such that disruption may manifest simultaneously as motor impairment and rigid, repetitive behavioral patterns.^15,16^

Approximately 10-20% of individuals with ASD have an identifiable genetic etiology, including rare variants in single genes or chromosomal abnormalities.^17-23^ High-throughput sequencing has identified over 250 genes that confer substantial ASD risk, including *SHANK3, DYRK1A*, and *SCN2A*, all of which are genes with established roles in synaptic function and neurodevelopment.^23-25^ These genetically attributable forms of ASD are increasingly recognized as having characteristic behavioral phenotypes that may differ from idiopathic ASD in ways beyond severity alone.^25-32^ For example, recent work has demonstrated that genetic variants can cluster into distinct phenotypic subgroups with different underlying biological programs, suggesting that genetic etiology may fundamentally shape the behavioral architecture of individuals with ASD.^5^ This work builds on earlier demonstrations that ASD polygenic risk scores are inversely associated with co-occurring developmental disabilities and that genetic architecture differs meaningfully across ASD presentations with and without intellectual disability.^33,34^ Despite this, most research examining motor-behavior associations has treated ASD as a unitary condition. This approach may obscure important differences in how behavioral domains relate to one another across genetic subtypes. If specific genetic variants disrupt shared neural circuits differently than the polygenic or environmental factors underlying idiopathic ASD, we might expect not only mean-level differences in motor and behavioral measures but also altered relationships between these domains.

This study examined whether the relationship between motor function and repetitive behaviors differs between children with ASD attributable to loss-of-function variants in high-confidence ASD risk genes (*SHANK3, DYRK1A*, and *SCN2A*), hereafter referred to as monogenic ASD, and children with idiopathic ASD. We hypothesized that the correlational structure between motor function and RRBs would differ between these groups, reflecting distinct patterns of neural circuit disruption. Using data from SPARK, we compared motor-RRB correlations between children with monogenic ASD and matched controls with idiopathic ASD and intellectual disability.^35^ We further examined whether any observed differences were specific to particular subtypes of repetitive behaviors and whether effects were consistent across genetic etiologies.

## METHODS

### Study Design and Participants

This study used data from SPARK, which collects genomic and phenotypic data from individuals with ASD and their families. The whole-exome sequencing dataset (iWES v3, August 2024) comprised of 142,128 samples.

Cases were children with ASD carrying loss-of-function variants in *SHANK3, DYRK1A, or SCN2A* (see Genetic Classification). Controls were children with ASD, intellectual disability (IQ <70), and no pathogenic variants in established ASD risk genes (FDR < 0.01 per Fu et al.^23^). This IQ restriction isolated gene-specific effects from cognitive confounds. Cases were matched 10:1 to controls on sex, age, area deprivation index, and genetic ancestry (first 5 principal components) using optimal pair matching when sufficient matches were available. The final sample included 93 monogenic cases and 787 matched controls.

### Measures

Motor function was assessed using the Developmental Coordination Disorder Questionnaire (DCDQ), a 15-item parent-report measure yielding 3 subscale scores: Control During Movement, Fine Motor/Handwriting, and General Coordination. Lower scores indicate greater impairment. A motor composite was calculated as the mean of the 3 subscales (α = 0.79).^36^

Restricted and repetitive behaviors were assessed using the Repetitive Behavior Scale-Revised (RBS-R), a 43-item measure yielding 6 subscales (α = 0.87).^37^ Following prior work^38,39^ we classified Stereotyped and Self-Injurious subscales as lower-order RRBs and Compulsive, Ritualistic, Sameness, and Restricted subscales as higher-order RRBs. Higher scores indicate greater severity.

### Genetic Classification

We analyzed rare (minor allele frequency <0.1% in both our dataset and the non-psychiatric subset of gnomAD v3.1) loss-of-function variants (protein-truncating variants, deletions, splice-site variants) in the following high-confidence ASD risk genes with sufficient case numbers for adequately powered analyses: *SHANK3, DYRK1A*, and *SCN2A*.^40^ Missense variants were excluded owing to uncertain functional consequences. Quality control using Hail version 0.2 excluded samples with outlying variant counts, low call rates, close relatedness, or sex mismatches. The final monogenic sample comprised 93 children (34 *SHANK3*, 46 *DYRK1A*, 13 *SCN2A*).

### Statistical Analysis

Spearman correlations between motor and RRB composites were calculated separately for monogenic and idiopathic groups. Group differences were tested using Fisher z transformation, with permutation testing (10,000 iterations) and bootstrap 95% CIs for robustness. We examined correlations with each RBS-R subscale separately, applying false discovery rate correction using the Benjamini-Hochberg procedure. To assess potential distributional artifacts, we compared coefficients of variation, calculated distributional overlap, restricted analyses to the common score range, and examined correlations within motor tertiles. Correlations were also calculated within each gene group and tested for heterogeneity using the Cochran Q statistic.

## RESULTS

### Sample Characteristics

The final analytic sample included 93 children with monogenic ASD (*SHANK3*, n = 34; *DYRK1A*, n = 46; *SCN2A*, n = 13) and 787 matched children with idiopathic ASD and intellectual disability. Groups were well-balanced on matching variables: mean age was 8.8 years (SD = 3.2) in monogenic cases and 8.9 years (SD = 3.1) in idiopathic ASD (p = 0.599), sex distribution did not differ between groups (p = 0.516), and IQ scores were also comparable between idiopathic controls (45.5; SD = 17.1) and monogenic cases (45.4; SD = 19.3). Sample characteristics are presented in Table 1.

**Table 1.** Sample Characteristics.

| **Characteristic** | **Idiopathic**<br>**ASD**<br>**(n = 787)** | **Monogenic**<br>**ASD**<br>**(n = 93)** | **P Value** |
| --- | --- | --- | --- |
| Age, mean (SD), y | 8.9 (3.1) | 8.8 (3.2) | .60 |
| Male sex, No. (%) | 604 (76.7) | 68 (73.1) | .52 |
| IQ, mean (SD) | 45.5 (17.1) | 45.4 (19.3) | .98 |
| Motor composite score (DCDQ), mean (SD) | 10.76 (3.94) | 8.05 (2.68) | <.001 |
| RRB total score, mean (SD) | 7.12 (4.1) | 5.27 (3.44) | <.001 |
| Higher-order RRB composite score, mean (SD) | 7.61 (4.63) | 5.50 (4.18) | <.001 |
| Lower-order RRB composite score, mean (SD) | 6.13 (4.14) | 4.79 (3.27) | .002 |

Abbreviations: ASD, autism spectrum disorder; DCDQ, Developmental Coordination Disorder Questionnaire; RRB, restricted and repetitive behavior.

Compared with idiopathic ASD, children with monogenic ASD showed significantly worse motor function (mean [SD], 8.05 [2.68] vs 10.76 [3.94]; P < 0.001) on the DCDQ. Monogenic cases also exhibited fewer repetitive behaviors on the RBS-R across both higher-order (mean [SD], 5.50 [4.18] vs 7.61 [4.63]; p < 0.001) and lower-order (mean [SD], 4.79 [3.27] vs 6.13 [4.14]; P = 0.002) domains. Despite these mean differences, all monogenic motor scores and all monogenic higher-order RRB scores fell within the observed idiopathic ASD range. A small proportion of idiopathic ASD cases fell outside the monogenic range, consistent with restricted-range analyses.

### Motor-RRB Correlations

The relationship between motor function and repetitive behaviors differed markedly between groups (Table 2). In idiopathic ASD, better motor function was associated with fewer repetitive behaviors (Spearman r = -0.156, P <.001). In monogenic ASD, the association was in the opposite direction (Spearman r = +0.185, P = 0.075). The between-group correlation difference was statistically significant (Δr = 0.341, Fisher’s z = 3.09, P =.002). For the higher-order composite specifically, permutation testing confirmed the group difference (P =.004), and the bootstrap 95% confidence interval for the correlation difference excluded zero (95% CI, 0.124-0.535).

**Table 2.** Motor Function-RRB Correlations by Group.

| **Outcome** | **Idiopathic**<br>**ASD**<br>***r*** | **Monogenic**<br>**ASD**<br>***r*** | **$\Delta r$** | **P Value** |
| --- | --- | --- | --- | --- |
| Total RRB composite | -0.156 | +0.185 | +0.341 | .002 |
| Higher-order RRB composite | -0.106 | +0.234 | +0.339 | .002<sup>a</sup> |
| Lower-order RRB composite | -0.246 | -0.033 | +0.212 | .05<sup>a</sup> |

Abbreviation: ASD, autism spectrum disorder; RRB, restricted and repetitive behavior.
Correlations are Spearman rank correlations. $\Delta r$ represents the difference between monogenic and idiopathic correlations.

### Specificity to Higher-Order RRBs

The correlation reversal was specific to higher-order repetitive behaviors (Table 2, Figure 1). For the higher-order composite (compulsive, ritualistic, sameness, and restricted behaviors), idiopathic ASD showed a weak negative association (r = -0.106, P = 0.003), whereas monogenic cases showed a positive association (r = +0.234, P = 0.024), representing a significant reversal (Δr = 0.339, P = 0.002). In contrast, the lower-order composite (stereotyped and self-injurious behaviors) showed attenuation but not reversal: idiopathic controls exhibited a negative association (r = -0.246, P <.001) that weakened in monogenic cases (r = -0.033, P = 0.751) but remained in the same direction (Δr = +0.212, P = 0.05).

**Figure 1.**
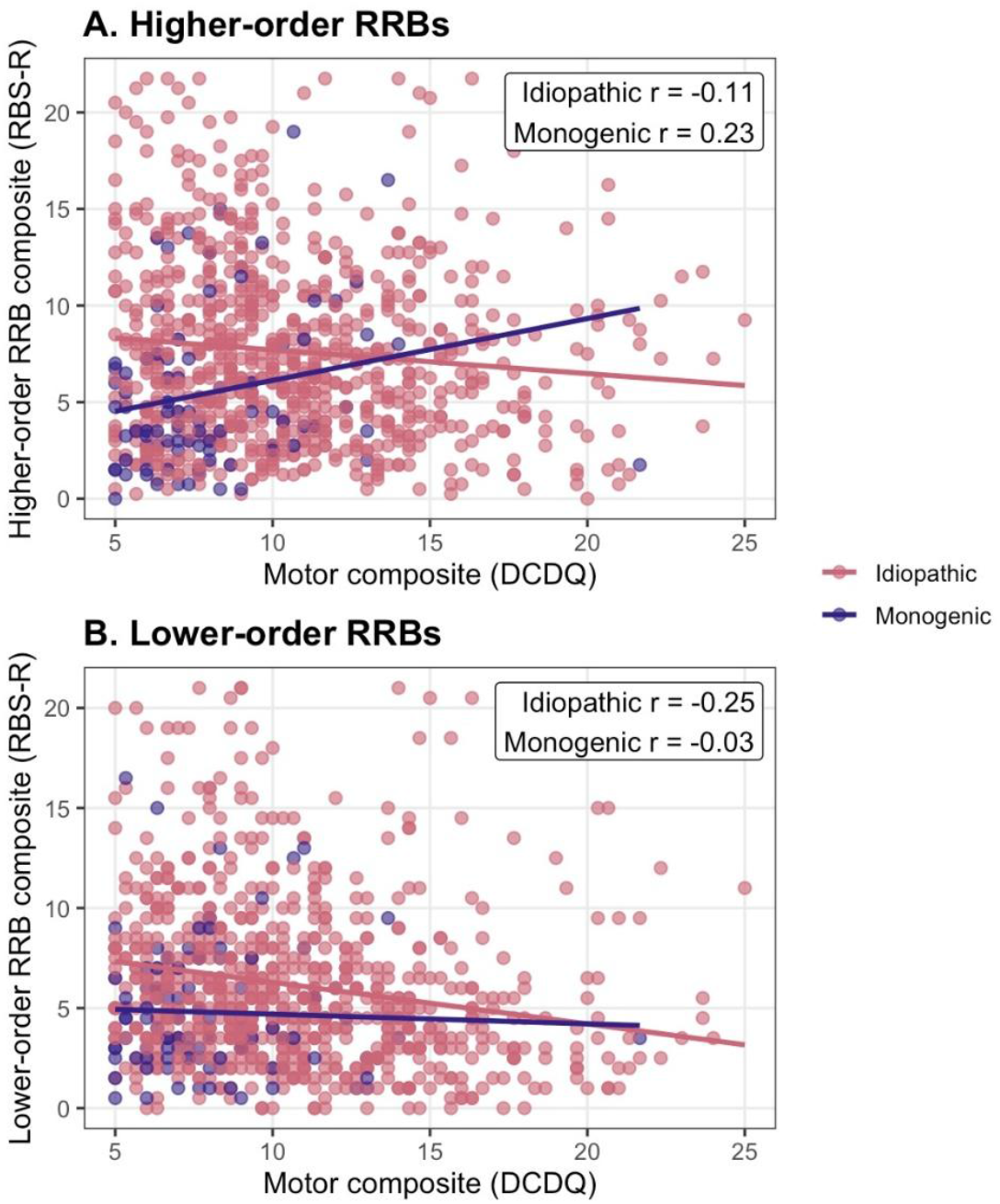
Motor Function-RRB Correlations by Higher and Lower Order.

Subscale-level analyses reinforced this pattern (Figure 2, Table S1). Three of four higher-order subscales showed significant correlation shifts after FDR correction: Compulsive (Δr = +0.386, P_FDR = 0.002), Ritualistic (Δr = +0.335, P_FDR = 0.006), and Sameness (Δr = +0.288, P_FDR = 0.018). The Restricted Interests subscale showed a similar but non-significant shift (Δr = +0.157, P_FDR = 0.183). Neither lower-order subscale reached significance: Stereotyped (Δr = +0.192, P_FDR = 0.115) and Self-Injurious (Δr = +0.146, P_FDR = 0.183). The mean correlation shift was larger for higher-order subscales (+0.292) than for lower-order subscales (+0.169).

**Figure 2.**
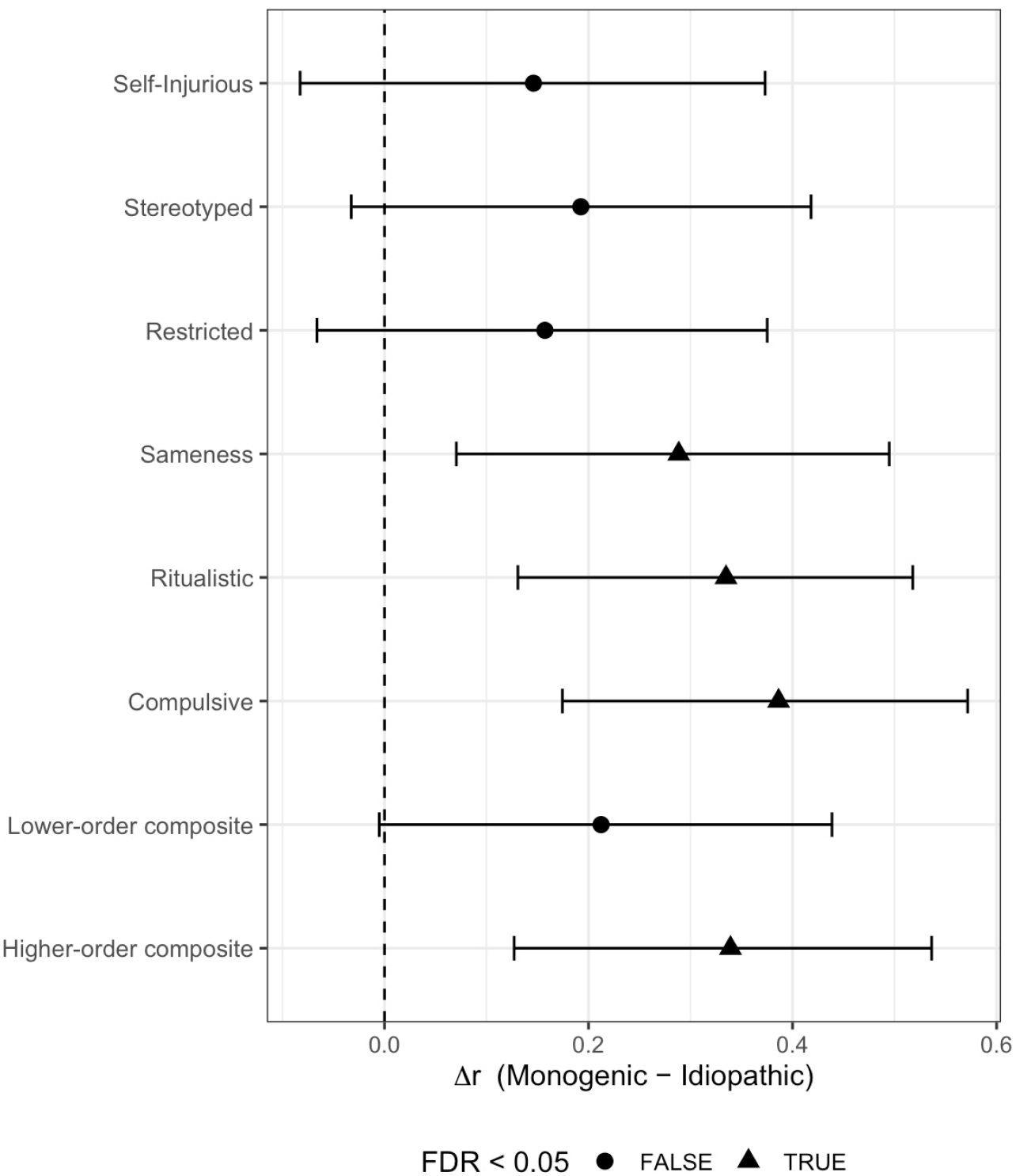
Motor Function-RRB Correlations by Higher and Lower Order Subscales.

### Sensitivity Analysis

We conducted several analyses to evaluate whether the observed higher-order motor-RRB correlation reversal could be attributed to distributional differences between idiopathic and monogenic groups (Table S2). Coefficients of variation were similar across groups for motor function (CV ratio, monogenic/idiopathic = 0.91) and were higher in monogenic cases for higher-order RRBs (CV ratio = 1.25), indicating that monogenic cases had adequate variability. When limiting both groups to the overlapping range of motor and higher-order RRB scores (retaining 96.6% of idiopathic controls (n = 760) and 100% of monogenic cases (n = 93), the reversal was maintained (idiopathic r = -0.104, monogenic r = +0.234, Δr = 0.338).

To determine whether the observed effect was driven by extreme motor scores, we examined motor-higher-order RRB correlations within motor strata defined using tertile cut points from the combined idiopathic and monogenic motor score distribution. As a result, group sizes within strata were unequal. In the low motor stratum (258 idiopathic controls; 65 monogenic cases), monogenic cases showed a more positive association than idiopathic controls (idiopathic r = +0.013, monogenic r = +0.136, Δr = 0.123). In the middle stratum (270 idiopathic controls; 19 monogenic cases), the difference was larger (idiopathic r = -0.154, monogenic r = +0.151, Δr = 0.305). The high motor stratum contained too few monogenic cases (n = 9) for reliable estimation. Together, these analyses indicate that the higher-order motor-RRB correlation reversal is not explained by mean differences, restricted range, or floor effects.

### Gene-Specific Patterns

To assess consistency across genetic etiologies, we examined motor-higher-order RRB correlations within each monogenic genetic subgroup (Table S3). All three genes showed positive correlations, in contrast to the negative correlation observed in idiopathic ASD: *SHANK3* (r = +0.033, n = 34), *DYRK1A* (r = +0.262, n = 46), and *SCN2A* (r = +0.623, n = 13). The test for heterogeneity was not significant (Cochran’s Q = 3.76, df = 2, P = 0.153), indicating no evidence that effect sizes differ substantially across genes given available sample sizes. The *SCN2A* group showed the strongest positive correlation, though this estimate should be interpreted cautiously given the small sample size (n = 13). *DYRK1A* showed a moderate positive correlation consistent with the pooled estimate. *SHANK3* showed a near-zero correlation that nonetheless represented a shift away from the negative association observed in idiopathic ASD.

## DISCUSSION

This study examined the relationship between motor function and repetitive behaviors in children with ASD who carry loss-of-function variants in *SHANK3, DYRK1A*, and *SCN2A*. Our findings suggest that the typical negative correlation between motor function and RRBs observed in idiopathic ASD reverses to a positive association in children with these genetic variants. This reversal was specific to higher-order repetitive behaviors such as compulsions, rituals, and insistence on sameness, and was not observed for lower-order sensorimotor behaviors. The effect was consistent across all three genes and remained robust across multiple sensitivity analyses.

The negative correlation we observed between motor function and RRBs among children with idiopathic ASD is consistent with findings in the broader ASD literature.^11-13^ Despite ASD having multiple symptom dimensions that may reflect independent genetic architectures, most genetic studies treat the disorder as a unitary phenotype. The emerging literature on ASD with identifiable genetic etiologies suggests that specific variants can produce distinct phenotypic profiles that differ from idiopathic ASD beyond symptom severity alone.^27,28^ ASD with an identifiable genetic etiology represents approximately 10-20% of cases, with monogenic syndromes accounting for roughly 5%.^29-32,41^ Comprehensive genetic evaluation can identify causative variants in up to 25-40% of individuals, particularly those with co-occurring intellectual disability or seizures.^42^ These genetically-defined forms of ASD are increasingly recognized as having characteristic behavioral phenotypes associated with specific genes.^18-21^ Our findings extend this work by demonstrating that genetic etiology can alter the correlational structure between behavioral domains, going beyond mean-level differences typically reported.

Although *SHANK3, DYRK1A*, and *SCN2A* affect different molecular pathways, our findings suggest all three may converge on shared mechanisms governing the motor-RRB relationship. Prior research has implicated frontal-striatal-cerebellar circuitry in both motor control and repetitive behaviors in ASD.^14-17^ Dysfunction in this circuitry has also been implicated in obsessive-compulsive disorder and Tourette syndrome, conditions characterized by repetitive behaviors and motor features that are notably comorbid with ASD.^43,44^ Each gene examined has been linked to this circuitry in preclinical studies: *SHANK3* encodes a synaptic scaffolding protein with high striatal expression, and mouse models demonstrate motor deficits alongside repetitive grooming^24,29,45^; *DYRK1A* haploinsufficiency is characterized by motor difficulties alongside stereotypic behaviors^26,31^; *SCN2A* loss-of-function mutations produce ASD phenotypes with motor impairments in mouse models.^25,32^ While our behavioral data cannot directly test circuit-level hypotheses, the convergence across these molecularly distinct genes is consistent with overlapping neural systems. This parallels the approach of Litman and colleagues (2025), who demonstrated that distinct phenotypic classes of autism are associated with different underlying genetic programs.^5^

The specificity of our findings to higher-order repetitive behaviors is notable. The distinction between lower-order motor stereotypies and higher-order cognitive repetitive behaviors is well-established and may reflect different underlying processes.^46,47^ Our finding that the correlation reversal was limited to higher-order behaviors suggests that the genetic variants examined may particularly affect behavioral flexibility and routine formation rather than basic motor patterns. This interpretation is consistent with prior work showing that early motor milestones predict later insistence-on-sameness behaviors specifically, suggesting a developmental link between motor function and cognitive repetitive behaviors.^39,48^

These findings may have clinical implications. Children with monogenic autism in our sample clustered at lower levels of both motor function and higher-order RRBs, with the positive correlation indicating that those with relatively preserved motor abilities showed more compulsions, rituals, and insistence on sameness. Given evidence that higher-order repetitive behaviors can serve self-regulatory functions,^46,47^ this pattern suggests these behaviors may require a threshold of motor and cognitive capacity to manifest. Children with the most profound impairments may lack the ability to deploy these complex regulatory strategies. Clinically, the emergence of higher-order RRBs as motor skills develop should not necessarily be interpreted as deterioration but may reflect acquisition of more sophisticated self-regulation. For children with monogenic autism, understanding the adaptive functions these behaviors serve, rather than targeting them uniformly for reduction, may be important. As interventions are validated for monogenic forms of autism, phenotypic patterns like the motor-repetitive behavior relationship could identify idiopathic subgroups who might respond to similar treatments, though prospective treatment studies are needed.

Several limitations warrant consideration. First, our sample sizes for individual genes were modest, particularly for *SCN2A* (n = 13), limiting statistical power for gene-specific analyses. Although the pooled analysis was adequately powered and the heterogeneity test suggested consistent effects, individual gene estimates should be interpreted cautiously. Second, both motor function and RRBs were assessed via parent report rather than direct observation. While the DCDQ and RBS-R are validated instruments, parent-report measures may be subject to shared method variance that could influence observed correlations. Third, our cross-sectional design precludes conclusions about developmental trajectories or causal relationships. Longitudinal studies examining whether motor-RRB relationships change over time would be valuable. Finally, our findings are specific to loss-of-function variants in three high-confidence ASD genes and may not generalize to other genetic etiologies or missense variants with different functional consequences.

## CONCLUSIONS

These results demonstrate that identifiable genetic etiology alters not only the severity of behavioral traits but also the correlational structure within developmental domains in ASD. The consistency of the correlation reversal across three genes with distinct molecular functions suggests shared underlying mechanisms. The finding that genetic status predicts different patterns of behavioral relationships supports the value of stratifying individuals with ASD by genetic etiology. As genetic testing becomes increasingly integrated into ASD clinical care, understanding how specific genetic etiologies shape behavioral phenotypes will be important for developing targeted interventions.

## Data Availability

The data used in this study are available through the Simons Foundation Powering Autism Research for Knowledge (SPARK) consortium. Qualified researchers may apply for access through SFARI Base (https://base.sfari.org).

## Acknowledgement

- *Concept and design:* Smout, Udeshi, Mahjani
- *Acquisition, analysis, or interpretation of data:* Caballero, Jung, Smout, Udeshi, Mahjani
- *Drafting of the manuscript:* Jung, Smout, Udeshi, Mahjani
- *Critical revision of the manuscript for important intellectual content:* All authors
- *Statistical analysis:* Smout, Udeshi
- *Obtained funding:* Mahjani
- *Supervision:* Mahjani

## Funding

The Seaver Foundation (Smout, Mahjani)

## Conflict of interest disclosures

None reported.

## Role of the funders

The funders of the study had no role in study design, data collection, data analysis, data interpretation, writing of the manuscript, or decision to submit the manuscript for publication.

## Notes

### Competing Interest Statement

The authors have declared no competing interest.

### Author Declarations

The Institutional Review Board of the Icahn School of Medicine at Mount Sinai gave ethical approval for this work.

