## Supplement for "Association Between Motor Function and Higher-Order Repetitive Behaviors in Monogenic Autism Spectrum Disorder"

**Supplemental Table 1.** Motor Function-RRB Correlations by Higher and Lower Order Subscales

| RRB Subscale | Idiopathic ASD *r* | Monogenic ASD *r* | Δ*r* | *P* Value[a] |
|---|---|---|---|---|
| **Higher-order** | | | | |
| Compulsive | ... | ... | +0.386 | .002 |
| Ritualistic | ... | ... | +0.335 | .006 |
| Sameness | ... | ... | +0.288 | .02 |
| Restricted interests | ... | ... | +0.157 | .18 |
| *Composite* | -0.106 | +0.234 | +0.339 | .002[b] |
| **Lower-order** | | | | |
| Stereotyped | ... | ... | +0.192 | .12 |
| Self-injurious | ... | ... | +0.146 | .18 |
| *Composite* | -0.246 | -0.033 | +0.212 | .05[b] |

Abbreviation: ASD, autism spectrum disorder; RRB, restricted and repetitive behavior.

[a] Unadjusted *P* value.

**Supplemental Table 2.** Sensitivity Analyses for Higher-Order Motor-RRB Correlations

| **Analysis** | **Idiopathic ASD** ***r*** | **Monogenic ASD** ***r*** | **Δ*r*** |
|---|---|---|---|
| Restricted range[a] | -0.104 | +0.234 | +0.338 |
| Low motor stratum (≤33rd percentile)[b] | +0.013 | +0.136 | +0.123 |
| Middle motor stratum (33rd-67th percentile)[c] | -0.154 | +0.151 | +0.305 |
| High motor stratum (≥67th percentile)[d] | ... | ... | ... |

Abbreviation: ASD, autism spectrum disorder; RRB, restricted and repetitive behavior.
[a] Includes 96.6% of idiopathic controls (n = 760) and 100% of monogenic cases (n = 93) with scores in the overlapping range.
[b] Includes 258 idiopathic controls and 65 monogenic cases.
[c] Includes 270 idiopathic controls and 19 monogenic cases.
[d] Insufficient monogenic sample size (n = 9) for reliable estimation.

**Supplemental Table 3.** Motor Function–Higher-Order RRB Correlations by Genetic Etiology

| **Group** | **No.** | ***r* (95% CI)** |
|---|---|---|
| Idiopathic ASD | 787 | -0.106 (-0.26, -0.11) |
| Monogenic ASD | | |
| *SHANK3* | 34 | +0.033 (-0.11, 0.03) |
| *DYRK1A* | 46 | +0.262 (-0.10, 0.26) |
| *SCN2A* | 13 | +0.623 (0.39, 0.62) |

Abbreviation: ASD, autism spectrum disorder; RRB, restricted and repetitive behavior.
Cochran Q test for heterogeneity: Q = 3.76, *df* = 2, *P* = .15.
